# Stakeholder views on implementing a novel addiction screening and prevention tool in a hospital setting: A qualitative study

**DOI:** 10.64898/2026.04.14.26350880

**Authors:** Genevieve F. Dash, Emily Balcke, Holly Poore, Danielle M. Dick

## Abstract

**Introduction:** Current best practice is for primary care physicians (PCPs) to screen patients for problematic substance use at checkups. However, this practice is not routine, is done in an unstandardized manner, and contributes to the overburdening of PCPs. Screening practices also target current, potentially problematic use behaviors, thus limiting their capacity to help patients prevent problems before they start. Recent scientific advances in identifying people at high risk for substance use problems as a means of facilitating prevention efforts have not yet been integrated into medical practice. To address these issues, our research team developed a freestanding platform called the Comprehensive Addiction Risk Evaluation System (CARES). CARES provides personalized information about genetic and behavioral/environmental risk for substance use disorder (SUD) and connects individuals to resources based on their risk profile. The present study evaluated the potential for adoption and implementation of CARES within a health care system through qualitative interviews with key stakeholders.

**Methods:** Semi-structured interviews were developed using the Consolidated Framework for Implementation Research (CFIR) and conducted with N=15 interviewees. Transcripts were analyzed using rapid qualitative analysis.

**Results:** Key themes included perceived need for new SUD screening tools, current SUD screening procedures and their pros/cons, openness to new ideas and clinical tools, fit of CARES with organizational goals and priorities, considerations for use of CARES with adolescent populations, anticipated patient response to CARES, barriers to implementation and uptake of CARES, changes required for implementation, and possibility for medical record integration. Interviewees generally expressed need for new screening tools and openness to using new tools, but expressed concern that existing provider burden, lack of SUD knowledge, and discomfort/stigma could stymie efforts to implement CARES.

**Conclusions:** There is a clear need for a low-burden, easy-to-use tool for substance use prevention. CARES appears to be an acceptable and feasible approach to fill this gap. These findings will be used to inform pilot implementation of CARES in a clinical care setting.

## 1. Introduction

Substance use disorders (SUDs) affect approximately 48 million individuals in the United States (Substance Abuse and Mental Health Services Administration, 2025). SUDs contribute to serious and significant downstream sequalae ranging from occupational and interpersonal problems to cancer and premature mortality (Stein, 1999; Weaver, 2010). Current screening practices are limited in scope insofar as they target current substance use and consequences, meaning that they are not ideally positioned to help patients prevent substance use problems *before* they start. A number of important scientific advancements have been made in understanding what variables predict an individual’s risk for SUD even before onset of problematic substance use behavior (Barr et al., 2022). In particular, at similar levels of use, factors such as behavioral traits (Pedersen et al., 2017; Smith & Cyders, 2016), current and past history of mental health disorders (King et al., 2004), sociodemographic variables (Castaldelli-Maia & Bhugra, 2022; Swendsen et al., 2009), and genetic risk (Deak & Johnson, 2021; Deak et al., 2019), may make some individuals more likely than others to develop SUD (Barr et al., 2022). Data from these multidisciplinary literatures represents a yet-untapped source of etiological information that can be incorporated into screening and early prevention/intervention of SUD.

Research on the structure of psychopathology has provided strong evidence for distinct but correlated dimensions of externalizing (e.g., impulsivity, risk-taking) and internalizing (e.g., anxiety, depression) that independently predict SUD (Dick et al., 2022). Because these behaviors often precede SUD onset (Farmer et al., 2015), and have well-validated and reliable measures, they are valuable sources of information on etiological risk for SUD readily quantified for application within risk screening contexts. There has also been research on the role of social determinants of SUD (Braveman et al., 2011; Lin et al., 2024), including neighborhood and working conditions, educational attainment, peer substance use, and childhood social disadvantage (Braveman et al., 2011; Lin et al., 2024). These factors are strong predictors of SUD (Lin et al., 2024), and can be used to guide quantification of environmental risk factors in SUD risk screening. Finally, advancement of psychiatric genetic methods has provided insight into the genetic mechanisms underlying SUD. For example, genome-wide polygenic scores (PGS) summarize effects of variants across the genome, providing a cumulative score of genetic risk for a particular phenotype in a given individual. With increasingly sophisticated methods and growing sample sizes, PGS now account for a significant proportion (∼10%) of the variance in psychiatric traits, including substance use and related externalizing outcomes (Karlsson Linnér et al., 2021). Further, PGS are significant predictors of substance use outcomes even after taking into account behavioral and environmental risk factors (Barr et al., 2022; Hung, in preparation). Increased accessibility of genotyping technology, including decreasing cost, make PGS increasingly feasible for integration into clinical contexts (Dick & Austin, 2026). Collectively, these research advances should allow us to identify at-risk individuals and prevent problems before they escalate, in line with the precision medicine goal of moving our health care system from one that is largely treatment-based, to one that is personalized, predictive, and preventative (Collins & Varmus, 2015).

Despite this, the healthcare system is faced with two major challenges that impede improvement in screening practices: 1) the high burden placed on medical providers (Quan et al., 2024), who are often responsible for screening for problematic substance use, and 2) recent scientific advances in identifying people at high risk for substance use problems have not yet been integrated into medical practice (Dick et al., 2022). Current best practice is for pediatricians and primary care physicians (PCPs) to screen patients for substance use at checkups (Babor et al., 2017; Babor et al., 2023; Hargraves et al., 2017). However, this practice is not routine, is done in an unstandardized manner, and contributes to the overburdening of PCPs (McNeely et al., 2021; Rosário et al., 2021). It can be particularly difficult for PCPs to decide, based on a patient’s brief description of their use, if the patient is engaging in truly problematic levels of use, if their use is causing impairment (an important criterion for SUD diagnosis), and which resources may be most appropriate based on their report (Blevins et al., 2018; Ross et al., 2015). Additionally, providers often report receiving limited training in addressing alcohol and drug use, leading to uncertainty about how to navigate a positive screen and lack of familiarity with treatment resources; as a result, many providers may avoid screening altogether (McNeely et al., 2018; Ross et al., 2015). Additionally, in part due to high demands placed on PCPs working to address several high-priority tasks within limited visit windows, comprehensive screening for behavioral, sociodemographic, and genetic risk factors is not presently included in standard assessment in pediatric or primary care settings. As a result, most patients are not currently benefiting from some of the most recent advances in the science of understanding risk for SUDs.

To address these concerns, our research team developed a freestanding platform called the Comprehensive Addiction Risk Evaluation System (CARES) (Dick et al., 2025). CARES provides a broad risk assessment based on participant responses to questions about behavioral and environmental risk factors that research has shown most robustly predict the development of problems (Barr et al., 2022), as well as genetic risk, generated through the provision of an at-home saliva sample from which DNA is extracted to create PGS. Participants are also screened for current levels of alcohol, cannabis, and tobacco use and are immediately connected to treatment resources if their responses indicate hazardous use. After providing personalized risk information, the platform provides participants with evidence-based resources designed to support their health and well-being. The goal of this program is to aid in 1) preventing the development of substance use problems in higher-risk individuals and/or 2) facilitating earlier intervention for individuals who have begun to use at problematic levels before their problems become severe. In this way, CARES has the potential to reduce provider burden and connect patients to resources informed by the most up-to-date research.

Members of our research team are currently conducting a randomized controlled trial of the CARES platform in a research setting (Dick et al., 2025). Initial results from N=188 participants show that a vast majority of participants understand the material presented within the CARES platform (95%) and agree that they learned new information in general (87%) and about genetic risk, specifically (82%). Most participants also reported that they enjoyed receiving their risk profile (92%) and felt that it was accurate (74%). Only 4% of participants reported feeling any degree of regret about their decision to receive genetic information.

Importantly, nearly all participants (95%) found the online delivery method to be convenient (Dick et al., 2025). Substance use outcomes following participation in CARES are forthcoming, but these preliminary data indicate that CARES represents a digestible, accessible platform that provides novel information to individuals interested in learning about their genetic and environmental risk for SUD. As a next step, and particularly as calls for translation of genetic research into clinical spaces grows (Dick & Austin, 2026), our team aimed to explore the viability of implementing CARES within a hospital-based patient care setting.

### 1.1 Present Study

The goal of the present study was to evaluate the potential for adoption and implementation of the CARES platform within a health care system through qualitative interviews with key stakeholders. To do this, we used an implementation mapping approach (Fernandez et al., 2019), which involves 5 key tasks: (1) conducting a needs assessment and identifying program adopters and implementers; (2) identifying performance objectives; (3) designing an implementation strategy; (4) producing implementation protocols and materials in collaboration with partner clinics; and (5) identifying implementation outcomes. This study presents the results of the needs assessment (Fernandez et al., 2019). Ultimately, the data collected here will be used to inform the launch of a CARES pilot in a selected clinic, following the protocol developed through the implementation mapping framework. We anticipate that conducting early implementation mapping and a small pilot of the implementation protocol will enhance the implementation outcomes of a larger evaluation of the CARES platform in a clinical setting.

## 2. Methods

### 2.1 Participants and Procedures

#### 2.1.1 Interviewee Recruitment

Collaborators within the healthcare system identified individuals for the needs assessment interviews. This was accomplished through word-of-mouth by leadership within the hospital system. System leadership selected providers within partner clinics to participate in interviews. Additionally, as part of the interview, participants were asked about other leaders, administrators, and/or providers who would be important for the research team to interview.

Potential participants were contacted via email. Inclusion criteria included being age 18 or over and a leader or provider within the university-affiliated healthcare systems. All procedures were approved by the Rutgers University Institutional Review Board.

#### 2.1.2 Interview Development

Interview development was guided by the 5 domains delineated by the Consolidated Framework for Implementation Research (CFIR) (Breimaier et al., 2015). These include: 1) intervention characteristics (e.g., need, capability), 2) outer setting (e.g., hospital system), 3) inner setting (e.g., hospital/clinic), 4) characteristics of individuals (e.g., leaders, facilitators, deliverers, recipients), and 5) process (e.g., engaging, adapting). The interview was developed iteratively by members of the research team and piloted ahead of data collection via a mock interview with members of the lab. Questions included in the final interview guide are available in the Supplemental Materials.

#### 2.1.3 Interview Protocol

Once a participant agreed to participate in an interview, they were provided with scheduling information, a consent form, and information about the CARES platform, including an overview of the program, images of the website, and sample feedback report (see Supplemental Material) via email. In-depth interviews in support of a needs assessment were conducted online via Zoom by at least two trained doctoral-level members of the research team who had clinical interviewing experience. Interviews were primarily facilitated by the first author (Dr. Dash) with support from a co-supervising author (Dr. Poore), who are both formally trained in structured and semi-structured interviewing; another co-supervising author (Dr. Dick) was also present for some of the interviews. Prior to beginning the interview, the interviewer reviewed the consent form and obtained verbal consent to proceed. Interviewees were also given the opportunity to review the CARES materials sent ahead of the interview and ask any outstanding questions about the platform to ensure adequate understanding of CARES. Interviews were 45-60 minutes and were recorded and transcribed within the Zoom application. Recordings and transcripts were only accessible to the research team. It was mandatory that the interview was recorded to ensure that the team was able to accurately analyze the response data following the interview and verify accuracy of the transcripts generated by Zoom. At the conclusion of the interview, recordings were immediately moved to a secure server in a folder only available to the research team and were deleted within the Zoom application. Interviewees were offered a $100 gift card for their participation. Interviews were conducted until little to no new information was generated, such that conducting further interviews would be unlikely to provide additional insight. The stopping point was agreed upon by 3 members of the research team. The final sample consisted of N=15 stakeholders from the healthcare system affiliated with Rutgers University, including employees at both specialty SUD and generalist clinics and institutes. The team’s consensus of saturation with this sample size is consistent with recommended sample sizes for reaching saturation in qualitative research (Hennink & Kaiser, 2022).

#### 2.1.4 Overview of CARES

An overview of CARES, including website images and a sample personalized feedback report, is available in the Supplemental Materials. The CARES platform can be openly accessed online (www.addictionrisk.com). CARES begins with a survey in which individuals provide demographic and contact information, as well as the CARES questionnaire of behavioral and environmental risk factors. Participants also provide a saliva sample. For community-based participants, a saliva sample kit is mailed with instructions for how to provide the sample and return the kit; for individuals in clinic settings, participants provide a saliva sample on site.

Individuals are notified via email when their results are available. Participants are then able to log into their CARES account and view their results. The CARES personalized feedback report provides an estimate of overall risk for SUD, which combines level of genetic and behavioral/environmental risk, and shows how many people with their profile go on to develop substance-related problems as compared to the national average. Risk estimates are also separated out into genetic and behavioral/environmental components. Additional information is provided about each of the components, including what environmental risk factors the individual carries and why those factors are associated with elevated risk, as well as education about how genes influence risk for SUD. The CARES platform was developed in collaboration with psychiatric genetic counselors and uses language intended to be empowering and destigmatizing (Dick et al., 2025).

### 2.2 Analytic Plan

Data were analyzed using rapid qualitative analysis, “an applied method used to obtain actionable, targeted qualitative data on a shorter timeline than traditional qualitative methods” (Lewinski et al., 2021). This approach was selected due to institutional demands of rapid turnaround of results ahead of implementation of the CARES pilot within an externally imposed timeline. Rapid qualitative analysis aims to identify intervention elements, key mechanisms, and program barriers in a time-sensitive, structured, and scientifically rigorous manner. There are 5 key steps: 1) create neutral domain names that correspond with interview questions, 2) create an interview transcript summary template guided by domains, 3) assess summary template usability and relevance, 4) summarize interview transcripts using the summary template, and 5) transfer summaries into a respondent-by-domain matrix to facilitate summarization of themes, domains, and respondent characteristics (Hamilton, 2013). Here, a descriptive hybrid approach was taken. Two doctoral-level members of the research team collaboratively and iteratively developed the domain names and templates, including updating templates for usability and accuracy following initial assessments of usability and relevance. All interviews were double-coded and any discrepancies were resolved through team discussion.

## 3. Results

### 3.1 Participant Characteristics

Participant characteristics are presented in Table 1. Most participants carried multiple roles and responsibilities across domains of leadership, patient care, administration, and research. All participants held a leadership role in some capacity, including executive leadership, institute and clinic directors and assistant directors, chief medical officers, and department chairs. A majority (75%) were patient-facing, approximately half (53%) held administrative roles, and a small but notable minority (20%) were actively involved in conducting their own research. Roughly half of participants’ positions involved work with SUD in some capacity (e.g., held a leadership position in a SUD-focused institute, worked as a clinician in an SUD specialty clinic).

**Table 1.** Participant characteristics.

| Gender | N (%) |
| --- | --- |
| Woman | 7 (47%) |
| Man | 8 (53%) |
| Role (Multiple Permitted) | N (%) |
| Leadership | 15 (100%) |
| Patient Care | 11 (75%) |
| Administration | 8 (53%) |
| Research | 3 (20%) |
| Highest Degree Earned | N (%) |
| MD | 7 (47%) |
| DO | 1 (7%) |
| MA | 3 (30%) |
| MSW | 1 (7%) |
| MPA/eMPA | 2 (13%) |
| MBA | 1 (7%) |
| SUD Focus in Current Position |  |
| Yes | 6 (40%) |
| Some | 2 (13%) |
| No | 7 (47%) |

### 3.2 Domains and Themes

Domains derived from conducting rapid qualitative analysis of the raw transcript data included perceived need for new SUD screening tools, current SUD screening procedures and their pros/cons, openness to new ideas and clinical tools, fit of CARES with organizational goals and priorities, considerations for use of CARES with adolescent populations, anticipated patient response to CARES, barriers to implementation and uptake of CARES, changes required for implementation, and possibility for medical record integration. Findings for each of these domains are described below in sections 3.2.1-3.2.5, categorized by the relevant CFIR domains (innovation characteristics, outer setting, inner setting, characteristics of individuals, process) under which they fall. There were three domains identified in (reimbursement practices, disseminating information about CARES to providers and clinics, key leaders) that are not described in results due to specificity to the healthcare system in which this implementation study is taking place and opting to prioritize focus on results more broadly relevant to implementation of a novel substance use screening tool like CARES. Some interviewee quotes have been edited minimally for clarity and readability, primarily to remove extraneous filler words. Care was taken to ensure that the meaning of each statement was accurately and adequately preserved.

#### 3.2.1 Innovation Characteristics

##### 3.2.1.1 Perceived Need for Updates to Screening (Relative Advantage of Innovation)

Interviewees were nearly universal in their perception that there is a need for new tools to screen for SUD in clinical spaces. Specifically, needs were identified for tools that are more individualized, promote value-based care, and facilitate prevention/early intervention in addition to screening for problems that may require treatment. Interviewees in several different roles observed how standard screening for biomedical and neurodevelopmental conditions has been accepted and integrated into routine practice, particularly for the purpose of early detection and intervention, but that such practices for SUD fall behind:

> “We screen for blood pressure. Why? Because there’s something we can do about it and save lives. So every time you go to primary care, you get a blood pressure check. Why? Because we care that we could catch something early. […] But our substance use disorder screening- and we’re a pretty mission-driven, advanced system- is terrible. So there’s nothing that exists, no one knows how to do it. We didn’t ever do it historically and we’re just working on bare minimum, even in the acute care setting.” (Physician, Emergency Medicine)
>
> “I think we’re missing substance abuse […] that if we had some form of screening tool, we would be able to intervene. I think parents probably would be craving it. I think, as parents, we want to do whatever we can to help our children early on, whether it’s assessment of autism, like we all talk about that- 2 years old, we want to get you in, assess, know if there is some autistic diagnosis so that we can start early intervention. Why is this any different than that?” (Administrator, Children’s Hospital)

Other reasons interviewees perceived a need for updates to existing tools were to more accurately capture use behaviors. Interviewees who work with adolescents, and college students in particular, noted that a provider simply asking about current use will typically elicit a “no,” regardless of actual use behavior, especially if the patient is presenting for an issue unrelated to substance use. It was also highlighted that most current screening practices focus on alcohol, nicotine, and cannabis use, which overlooks other substances that patients may be using.

#### 3.2.2 Outer Setting

##### 3.2.2.1 Current Screening Procedures and Their Pros/Cons (Local Attitudes, Policies)

Current practices within the university healthcare system were highly variable across settings and specialties, with no standardized, system-wide practice or guideline in place. SUD screening and evaluation processes were more structured and defined in SUD and emergency department settings, but much less so in primary care settings. In SUD-focused settings, it was common for the Tobacco, Alcohol, Prescription medication, and other Substance use tool (TAPS) (Wu et al., 2016) to be universally administered, as well as an American Society of Addiction Medicine (ASAM) assessment for patients interested in SUD treatment services (Mee-Lee et al., 2013); in some clinics, Narcan was given to patients take home. For programs more focused on prevention and intervention, the Alcohol Use Disorders Identification Test (AUDIT) (Reinert & Allen, 2002) and Cannabis Use Disorder Identification Test (CUDIT) (Adamson et al., 2010) were administered as standard protocol. Interviewees working in ED settings reported universal screening questions about current substance use during triage, with follow-up from a peer recovery specialist if a patient screened positive for substance use risk. Primary care providers and family physicians reported no formal screening procedures and inconsistent screening practices, in some cases conducted via informal observation; however, some did report training and occasional use of tools such as the AUDIT and CAGE questions (“Cut down, Annoyed, Guilty, and Eye-opener”). There were several reasons for the inconsistent screening practices observed in many clinics, including competing demands and provider burden, uncertainty about what resources would be available in the event that screening suggests need for intervention, and perceived patient receptivity to talking about substance use. One interviewee captures these interwoven challenges:

> “I think just competing demands […] we see a lot of people with tons of chronic conditions, and so in our 20-minute visit, we’re usually tackling already 4 or 5 problems at a time. And there’s also challenges of everybody wanting us to do more and more and more […] The second [challenge] is probably not having good treatment resources, so I think some people say, well, you know, if we screen for somebody and they screen positive, now what happens? Where do we send them? […] And then the third thing is probably patient receptivity. Some patients don’t want to talk about it.” (Physician and Researcher, Family Medicine Clinic)

These issues were reflected by other interviewees to varying degrees.

> “I think the major problem that they [providers] have is that they don’t know what to do with them [patients] if this [screen] came positive. So, very often they would shy away from doing even the most basic screening. I’m thinking about the primary care outpatient services that we have. They’re just so overwhelmed and they do not have the manpower, womanpower to provide substance use treatment, so I think that the screening is minimal.” (Physician and Leadership, Specialty Clinic)

A family physician highlighted how familiarity with, confidence in, and availability of treatment resources impacts screening. Notably, this varied across substances.

> “In primary care, we screen for stuff we think we can help treat, right? So, I think that talking to people and screening for opioid use disorder- because we have a really good treatment for opioid use disorder that we can give, that we feel really comfortable with-feels easier and more natural […] I think where we struggle more is with alcohol […] Most clinicians in my practice are not as comfortable with treatment of alcohol use disorder.” (Physician and Leadership, Family Medicine Clinic)

As evidenced in these responses, interviewees associated screening protocols with identifying current problems or disorder and referring to treatment. In almost no case did interviewees identify screening as a potential tool for prevention unprompted. Given that the lack of knowledge, accessibility, and access to treatment resources were primary barriers to utilizing screening protocols, a preventive approach such as CARES may help to facilitate uptake by circumventing the burden placed on providers when a positive screen is expected to be followed by connection to treatment resources.

#### 3.2.3 Inner Setting

##### 3.2.3.1 Openness to New Ideas and Clinical Tools (Culture, Tension for Change)

Nearly every interviewee expressed that they were open to the idea of CARES and to introducing a new screening tool for SUD. Most interviewees also indicated that they believed their colleagues would be open to CARES. One interviewee observed that this may be in part due to the nature of being part of an academic healthcare system that values evidence-based medicine and implementation of new tools grounded in up-to-date biomedical research:

> “I know that my group of physicians loves evidence-based medicine and loves using cool new tools […] they love doing new stuff if it really shows that there could be a benefit. They love doing it. They *don’t* like doing things because it comes from the top down.” (Physician, Pediatrics Clinic)

However, most interviewees also qualified this openness with considering how integrating CARES into the clinic may add to provider burden and impact workflow:

> “It just comes down to the time commitment…everybody’s so busy, they don’t want do one more thing.” (Physician and Researcher, Family Medicine Clinic)
>
> “If it’s presented in a manner that doesn’t add another thing to [providers’] plate- and not just them, their team […] They’ll be more open if it’s easy.” (Administrator, Pediatrics)
>
> “It’s really just about whether or not it’s going to be easy.” (Physician, Emergency Medicine)

Some interviewees noted a high likelihood of resistance, attributing this primarily to difficulties with change and asking staff to modify workflow. One interviewee also noted that stigma around SUD may impact openness, also returning to potential burden on providers.

> “I don’t think it’s the hesitancy around integrating research opportunities into care delivery, I think it’s more the stigma […] I think the other consideration that’s really important is there’s obviously a time demand on the provider.” (Leadership, SUD-Focused Institute)

Overall, openness to the idea of CARES was high. However, interviewees consistently noted a disconnect between openness to this new tool in theory versus in practice, with a heavy focus on additional provider burden, ease of integration into workflow, and usability.

##### 3.2.3.2 Fit of CARES with Organizational Goals and Priorities (Mission Alignment)

Interviewee’s perceptions of how well CARES would fit with their clinic or institute’s goals and priorities, as expected, varied by the clinic or institute’s focus. Primary care and family physicians generally prioritized screening for other biomedical conditions, with substance use and related problems being less of a priority:

> “The top [conditions to screen for] are the cardiovascular ones, diabetes, high cholesterol, high blood pressure, people with heart disease, and then cancers. After that, then it becomes more the respiratory ones, COPDs, asthma. That’s the majority of our patients…I would say substance abuse [problems] are lower down on there.” (Physician and Researcher, Family Medicine Clinic)

Other interviewees noted nuance regarding relative priority of CARES based on clinic focus, presenting concerns of the patient, and broader environmental factors:

> “Maybe it’s in the patients who have already moved into the behavioral health arena that there may be greater receptivity as an additional tool for a psychologist or social worker.” (Leadership, SUD-Focused Institute)
>
> “I don’t necessarily see [CARES] being not a priority. I think this time of year is tough with the new year…people are coming into the ED with all kinds of flu, COVID, and all that. So I would say, if you’re looking to implement it, I would say the spring or the fall.” (Leadership, SUD Clinic)

Several interviewees noted that CARES fits with goals focused on harm reduction and the shift that healthcare more broadly has been making away from abstinence-based programming:

> “As a priority, I think it’s a great time, because the focus is really shifting to harm reduction, and part of harm reduction is the prevention part. So, to me, [CARES] is a nice fit.” (Leadership, SUD Clinic)
>
> “That’s part of a harm reduction focus, which is giving people information to make good decisions going forward. That, to me, is essential.” (Leadership and Clinician, University Health Center)
>
> “The numbers of patients that don’t want recovery and abstinence, we are not serving. We’re sending them out into the street to either survive or not survive. So, we are really committed to harm reduction, which is going to change how we do business.” (Leadership, SUD Clinic)

Unsurprisingly, the perceived fit of CARES with organizational goals and priorities tended to be higher among interviewees with SUD-specific expertise and/or who work in the SUD space.

Physicians and providers working with a more general population emphasized that other biomedical conditions that contribute to high rates of morbidity and mortality, and for which clearer and more accessible intervention protocols are available, take priority over screening for SUD.

#### 3.2.4 Characteristics of Individuals

##### 3.2.4.1 Considerations for Using CARES with Adolescents (Innovation Recipients)

(Merikangas & McClair, 2012)Interviewees noted that parents of adolescents would be particularly interested in participating in a program like CARES (“This is what you do for kids. They love to find out about themselves” [Leadership, SUD Clinic]). However, several challenges were also noted. These included ethical considerations for providing services to individuals under age 18, including implications for parental involvement and privacy of the adolescent. Some interviewees also noted that parents may decline participation for their adolescent because they do not think it is relevant to their child:

> “When I talk about harm reduction in adolescents, it has to be a little harsh. People are just kind of laissez-faire about this. They don’t think it’s their kid. They don’t think it’s going to be their problem.” (Physician, Emergency Medicine)

One interviewee also noted that exploratory substance use is, to a degree, normative in this age group, and that risk feedback at this stage may “create a lot of anxiety for everyone, as kids kind of go through the normal exploratory [phase] in adulthood” (Physician and Leadership, Family Medicine Clinic).

#### 3.2.5 Process

##### 3.2.5.1 Anticipated Patient Response to CARES (Assessing Needs- Innovation Recipients)

Anticipated patient response to CARES varied by clinic and target population. Unsurprisingly, interviewees who worked in SUD specialty care generally perceived a more positive patient response to CARES:

> “Maybe a good thing that could come of this is so many [patients] are just like, whoa…I’m getting some wild information here. Maybe I need to talk to someone.” (Leadership and Clinician, University Health Center)
>
> “There’s no stigma. It’s not judgmental, and it’s not my fault. I’m not responsible for my disease, I’m responsible for my recovery.” (Leadership, SUD Clinic)

In contrast, physicians who work with a broad range of presenting concerns, and particularly with older adults, did not see CARES as being particularly relevant or of interest to their patients:

> “When I think about my patient population, it’s hard for me to imagine many people would be interested in [CARES].” (Physician and Leadership, Family Medicine Clinic)

This was attributed to believing that older adults have more pressing, acute health concerns, including heart conditions and chronic pain, and are outside of the window of risk for development of SUD (adolescence to young adulthood).

##### 3.2.5.2 Patient Barriers to Implementation and Uptake of CARES (Assessing Needs- Innovation Recipients)

The most commonly cited patient barrier to participating in CARES was potential response to receiving risk feedback. In particular, interviewees mentioned patients not wanting to know about their risk or parents of adolescents who may participate in CARES not wanting to “admit” that substance-related problems might occur in their family or household. Stigma was also mentioned in this context, insofar as patients may prefer to not know their risk rather than have a “scarlet letter” (Administrator, Children’s Hospital). Similarly, one interviewee observed a common patient preference to keep SUD treatment and care separate from their other medical care, in part to avoid having their presence in SUD treatment potentially result in stigmatization by their PCP and, in turn, negatively impacting their medical care. Two providers also noted historical mistrust of medical research and the medical establishment more broadly among minoritized groups, and particularly the Black community, with these groups being “rightfully wary of novel things that scientists want to do” (Physician, Emergency Medicine) and holding skepticism of genetic testing in particular.

##### 3.2.5.3 Provider Barriers to Implementation and Uptake of CARES (Assessing Needs- Innovation Deliverers)

Consistent with issues impacting provider *openness*, the primary barriers to provider *uptake* of CARES were provider burden and associated challenges. High demand on providers, need to address several presenting concerns within limited visit windows, and prioritization of other conditions (e.g., cancer) were mentioned as elements of this.

> “One of our most precious resources is time, right? We always have to choose: what’s the most important thing to address? So, it’s hard for me to imagine incorporating this as a regular thing as a routine part of our process.” (Physician and Leadership, Family Medicine Clinic)

Liability issues were also mentioned, including unclear scope of responsibility with respect to information provided by the CARES platform.

> “From the clinician’s perspective, I think we have a major issue with liability. And I’m not talking about getting sued, although that includes that part. But more about, what’s my scope of responsibility here? If I get a report that shows a high risk for impulsivity, depression, and suicidality, high risk for X, Y, and Z, even if the app or the system gives you some resources, ultimately I’m the quarterback. I am the one who’s going to organize all that. And it’s in my progress notes that I used that tool and that tool showed an increased risk. […] What did you do about that? How did you follow up?” (Physician and Leadership, Specialty Clinic)

Interviewees also broadly described provider discomfort with talking about substance use. There were several reasons for this. Interviewees mentioned that providers may need additional training, knowledge, and qualifications to address issues that patients might bring up if providers open the door to talking about substance use; similarly, providers may feel as though they would need to track and maintain more information about next steps for patients who want or need additional resources. Similarly, interviewees noted providers being concerned about not having expertise in genetics and being unsure about how to discuss such results with their patients, should their patients bring in their CARES results and wish to discuss them with their provider. Several interviewees also mentioned individual provider practice styles and preferences, noting that some providers prefer not to change how they approach their practice, that providers do not like to bring materials outside of their expertise into their practice, and that CARES may be perceived as “disrespecting the clinical acumen” of providers.

> “You will find that different personalities matter more. That there are some people who are just like, “Listen, I’m not doing that nonsense. This is how I’ve practiced forever.” (Physician, Pediatrics Clinic)

However, overall, the issues of time limits and additional burden were the most salient and consistently reported concerns across interviewees.

##### 3.2.5.4 Organizational/Administrative Barriers to Implementation and Uptake of CARES (Assessing Context)

Primary barriers at the organizational/administrative level primarily surrounded difficulty of enacting system-wide implementation (“getting a system to buy into something, that’s a challenge” [Senior Leadership, SUD-Focused Institute, Female]). One interviewee noted how implementation may be more successful in individual primary care clinics, as opposed to hospital- or system-wide change. This was, in part, due to competing demands at the hospital-level:

> “I think primary care offices will be the easiest, because they are very used to having resources, getting those resources out, promoting those resources within the office…The hospitals are a little more difficult…it’s like trying to kind of fight to be that priority. And the ED, it is tough. It is very tough to get any attention on things but primary care. Without a doubt, I think it would be a great tool.” (Leadership, SUD Clinic).

Other barriers included limited staffing, staff burnout with navigating many changes in the healthcare system, and increasing screening potentially “open[ing] the floodgates” (Administrator, Children’s Hospital) of treatment need within an already strained system.

##### 3.2.5.5 Suggested Changes to Organization/Clinic Ahead of Implementation (Tailoring Strategies)

Two primary needs within organizations/clinics were consistently raised by interviewees. The first was achieving “buy in” from leadership and providers. One interviewee noted the need to “convince local physicians first, because you want ground support” (Leadership, Internal Medicine) another stated: “Provider buy in is the first step” (Leadership, SUD Clinic). Obtaining CEO approval was also noted as potentially the easiest way to facilitate uptake of CARES, but that this was unlikely. The second change was to integrate CARES materials into clinics, making information accessible to providers and patients in multiple places around the clinic to facilitate exposure. Suggestions included stocking pamphlets in the waiting room and identifying a program champion to help facilitate this process. One interviewee also mentioned the potential for offering continuing education credits for obtaining training in CARES.

##### 3.2.5.6 Suggested Changes to CARES Ahead of Implementation (Reflecting & Evaluating, Tailoring Strategies, Adapting)

In preparation for piloting CARES in a clinical setting, interviewees were asked about any concrete changes they believe could be made to the platform to enhance uptake and make it a better fit for their patient population. Three interviewees suggested modifying the platform to make it more accessible by reducing the amount of text, making the text more “digestible” to a range of reading levels (CARES is currently written for a high-school reading level), and translating the platform to Spanish. Other related suggestions included making the website more accessible to individuals with reading or vision difficulties by providing videos or audio recordings. Two interviewees also mentioned that removing the word “addiction” from the platform name may increase interest and reach. Four interviewees recommended providing follow-up resources, such as an option to have results forwarded to the participants’ provider or the ability to speak with a genetic counselor connected with the CARES platform to discuss results. With respect to longer-term changes, three interviewees described the importance of having evidence of CARES’s long-term efficacy to make clear what clinics, providers, and patients will gain by using the platform (“Time is the precious resource, so every screening we do has to be evidence-based. We have to have a really robust reason for doing stuff” [Physician and Leadership, Family Medicine Clinic]).

##### 3.2.5.7 Medical Record Integration (Tailoring Strategies)

Interviewee’s perspectives on integrating CARES into the hospital system’s medical record were mixed, although a majority of interviewees expressed preference for keeping CARES separate from the medical record (“I would totally want to keep it freestanding” [Physician and Leadership, Family Medicine Clinic). Noted benefits of integration included ability to use the electronic medical record system to disseminate CARES, benefits for workflow, and ability for providers to track and follow up with patients who are high risk. Reasons for keeping CARES separate from the medical records were concerns about patient privacy, potential for stigmatization if high-risk results were in the chart (e.g., could impact future prescribing choices), that medical record integration would ultimately create more provider burden, and feasibility such that that the barriers to integration at the systemic and administrative levels are very high. Some interviewees also noted that having CARES be private and separate from the medical record might increase feelings of patient autonomy (“the more privacy I have, the more I own it as a tool for me” [Leadership, SUD Clinic]).

## 4. Discussion

The present study evaluated the potential for adoption and implementation of CARES, a freestanding online platform designed to provide individuals with personalized information about their risk for SUD (Dick et al., 2025), in a clinical care setting. This study represents step one (conducting a needs assessment and identifying program adopters and implementers) of the implementation mapping approach taken here, guided by the Consolidated Framework for Implementation Research (CFIR). Broadly, interviewees saw significant limitations with current SUD screening protocols and an associated need for novel screening tools, particularly ones that reduce provider burden and promote individualized patient care. Lack of familiarity with SUD screening and follow-up, general discomfort with discussing SUD, and burden of implementing a new tool were identified as counterbalancing a general openness to uptake of CARES. Fit of CARES with organizational goals and perceived patient interest in CARES varied based on clinic and patient population. In particular, SUD-focused providers, pediatricians, and ER providers saw CARES as a good fit within their practice. Primary care physicians were variable in their responses, with physicians working with younger populations seeing potential utility and physicians working with primarily older populations being unsure as to the value of CARES; risk for development of SUD peaks during adolescence and young adulthood (Merikangas & McClair, 2012), such that CARES may be most impactful among younger age groups. A consistent and recurrent theme was the need for a screening tool that is easy for providers to use, takes minimal time and training to implement, and primarily targets a younger population (adolescents and young adults). Interestingly, rather than creating an ease of workflow, many interviewees saw integrating the tool into the medical record as potentially introducing additional burden and liability for providers, and a majority of interviewees recommended keeping the platform freestanding. Although themes of provider burden and lack of familiarity have been identified in past studies of barriers to and facilitators of SUD screening (McNeely et al., 2018), this is, to our knowledge, the first study evaluating stakeholder views on a genetically-informed substance use screening and prevention tool for application in clinical settings.

The clear need for a low-burden, easy-to-use tool, coupled with the lack of consistency in screening protocols across clinics within the healthcare system, creates an ideal opportunity for implementation of CARES, which was designed to be patient-driven, to be minimally reliant on provider involvement, and to require no training or specialized knowledge about SUD to effectively use and benefit from. The nature of CARES positions providers to be disseminators of this screening tool, rather than administrators, actively reducing provider burden in the context of SUD screening; this is further bolstered by keeping CARES separate from the medical record, which serves to minimize provider burden, reduce patient concerns about stigma and empower patients to make informed and self-directed choices about substance use based on information that is maximally tailored to their individual risk and current levels of substance use. Interviewee feedback suggested that there is both a need for a tool like CARES within the hospital system, and that implementation of such a tool within some of the system’s clinics is a feasible approach to addressing current limitations of SUD screening.

Results of this needs assessment also provided insight into important modifications to the CARES platform to make it maximally useful to a broad audience and to facilitate the implementation process. In response to feedback provided by interviewees, we implemented the following changes to CARES: 1) created a Spanish language version of the CARES platform and translated all resources into Spanish, 2) created additional handouts to more easily connect patients to local and online resources related to substance use and mental health, 3) created handouts with conversation starters to facilitate discussion about risk assessment results between patients and their families and providers, and 4) offered access to a genetic counselor with whom patients could consult regarding their results. Based on the results of this needs assessment, our team is carrying out a small pilot to enact the implementation strategy resulting from this feedback. We have partnered with a pediatric emergency room department to recruit patients to complete CARES in the healthcare setting. As part of the implementation study, we are conducting a series of surveys with the patients to assess their satisfaction with the program, perception of its usefulness, any behavior change that resulted from receiving their addiction risk results, and the extent to which they used the resources provided. We are also soliciting feedback from providers regarding their perception of the platform’s usefulness in screening for addiction risk and ease of use.

### 4.1 Limitations

The findings summarized here represent a critical first step in implementing truly translational intervention into clinical care for patients at risk for SUD. However, several limitations should be considered. First, these findings are localized to a single healthcare system with an academic connection. The viewpoints expressed by interviewees are undoubtedly shaped by the culture of the healthcare system, and may thus represent a more receptive audience to a program such as CARES. Second, due to the rapid turnaround often necessitated in implementation settings and clinical contexts, we opted to utilize rapid qualitative analyses rather than a more intensive analytic approach, such as thematic analysis. While a more pragmatic approach within the parameters of the study and implementation timeline, this approach may not have yielded the depth of findings that another analytic approach might have achieved. Finally, we did not conduct a formal member checking process.

### 4.2 Conclusions

There is clear need for novel approaches to SUD screening that address current limitations with existing protocols, including focus on existing problems (thereby limiting potential for prevention of problems before they start), high provider burden resulting in inconsistent and, at times, ineffective screening, and lack of integration of recent scientific advancements in identification of individuals at high risk for SUD into routine medical practice. CARES aims to address these gaps by providing physicians and patients with a freestanding platform that provides an individualized risk assessment to patients regardless of level of current substance use involvement. Overall, the idea of implementing CARES was well-received by key stakeholders, despite a handful of notable barriers to be considered in the implementation process. Our team’s on-going pilot implementing CARES in a pediatric emergency department setting will provide insight into the feasibility and early efficacy of the use of such a tool for identifying substance use risk and preventing hazardous substance use. The positive receipt of CARES by providers in SUD clinics, EDS, and- depending on target patient population- primary care clinics, provides valuable insight into the potential reach of uptake and utility of CARES. Results of the pilot will help to guide next steps in implementation and dissemination of this novel SUD screening tool.

## Author Note

This work was funded by grant number P30HS029759, Agency for Healthcare Research and Quality (AHRQ).

## Author contributions

GFD- Methodology, formal analysis, investigation, data curation, writing-original draft, writing- review and editing; EB- Writing- review and editing, project administration; HP- Conceptualization, methodology, formal analysis, investigation, data curation, writing-review and editing, supervision, funding acquisition; DMD- Conceptualization, investigation, resources, writing- review and editing, supervision, funding acquisition.

## Data Availability

To preserve participant privacy, data will not be made publicly available.

