## Supplemental Materials for "Stakeholder views on implementing a novel addiction screening and prevention tool in a hospital setting: A qualitative study"

**for**

### **In-Depth Interview Guide**

### **Aim 1, Needs Assessment Interviews**

**Introduction**

To start, tell me a bit about your role in [clinic, department].

1. What is your profession?
2. How long have you worked in health care?
3. What types of conditions do you treat?

**Culture**

1. First I want to ask some broad questions about your view on the division/clinic’s culture around integrating new scientific developments and research findings into clinical care.
   - What are people’s general responses to participating in research or being guided to integrate new research findings into their practice?
   - To what extent are new ideas embraced and used to make improvements in your [clinic, department]?

**Compatibility** What makes a new tool/program a good fit for your clinic?

1. How well do you think CARES in particular could fit with existing work processes and practices in your setting?
   - What are likely issues or complications that may arise?

**Tension for Change**

1. Is there is a current screening process in place to evaluate risk for substance use problems in your division/clinic/department?
2. What are the current approaches used at the [Division, Clinic or Department] for screening patients for substance use problems?
3. What types of challenges do you think clinicians face with screening patients for substance use problems?
4. Do you see a need at the provider and patient levels for a tool like CARES in your clinic? Why or why not?
5. Do you think your providers/staff see a need for a resource like CARES? Tell me more.
6. How do you think providers will feel about being asked to integrate a new tool into their practice?

**Networks & Communications**

1. What would be the most helpful way for people in your division/clinic to receive information about a new initiative like CARES (e.g., email, all staff meeting, training, brochure/flyer)?
2. How do providers in your clinic typically receive information about training for use of a new tool? What would it take to reach providers and provide them with training/information about using CARES?

**Structural Characteristics**

1. The CARES platform has a cost associated with it, about $150 per patient to run the genotypes and administer the website. We have grant funding to cover the costs during our implementation trial so we can provide CARES at no cost to the clinics. We are also thinking long-term about how integrating CARES into clinical care might look from a billing and financial perspective.
   a. How are SUD services usually covered (e.g., the peer recovery services)?
   b. Is an evidence-based assessment and intervention platform like CARES something that could be reimbursed by insurance?
   c. If insurance reimbursement isn’t feasible, what would it take for the hospital to be willing to cover the cost of CARES?
2. What are important aspects of the [Division, Clinic, Department] infrastructure (social architecture, age, maturity, size, or physical layout) that would be important to consider in the process of introducing and implementing CARES?
   - What kinds of things might make implementation of CARES challenging; are there any particular barriers that would be important for us to know about?
3. Are there any infrastructure changes that would be needed to accommodate CARES?
   - Changes in scope of practice? Changes in formal policies? Changes in information systems or electronic records systems? Other?
   - What kind of approvals will be needed? Who will need to be involved?
   - Can you describe the process that will be needed to make these changes?
4. CARES currently operates as a freestanding platform. Do you think patients would be interested in having the platform integrated with their medical record or prefer to keep it separate?

As a provider, what is your perspective on the value of medical record integration in this context?

- Would you want the patient to have the option to also have the results of their addiction risk evaluation sent to you? Why or why not?

**Patient Needs & Resources**

1. How do you think patients will respond to CARES?
2. What barriers will patients face to participating in CARES?
3. How well do you think CARES will meet the needs of patients?
   a. In what ways could CARES meet their needs? E.g. improved access to services? Help with self-management?
4. [*For interviewees in pediatrics/adolescent care contexts*]: CARES is written at a high school level and applicable to individuals who are approximately 14-15 years of age or older.
   a. How does CARES compare to your current standard for screening for substance use in adolescents?
      i. Do you think CARES would be a useful tool for adolescents? Why or why not?
   b. Do you think your adolescent patients might be more forthcoming about substance use on a platform like CARES compared to disclosing to their provider directly?
      i. Is this a pro or con of the platform? (In other words, are there concerns about adolescents providing this information on a platform that the provider cannot access?)
   c. What other questions or concerns would you have about using the platform with minors?
   d. Would you limit use to individuals age 18 and older? Why or why not?
   e. Are there implementation considerations for using CARES with adolescents?
      i. Where do you see parents fitting into the equation?

**Goals & Feedback**

1. How could implementation of CARES align with organizational goals in your division/clinic?

**Relative Priority**

1. What kinds of high-priority initiatives or activities are already happening in your setting?
   - What is the priority of getting CARES implemented relative to other initiatives that are happening now?
     1. If not very high, what kinds of support and additional resources might help to facilitate implementation?
   - Will the implementation of CARES conflict with these priorities? If yes, how?
   - Will the implementation of CARES help achieve (or relieve pressure related to) these priorities? Why or why not?

**Engaging** **Opinion Leaders**

1. Who are the key influential individuals to get on board with this implementation?
